# Associations Between Exercise Capacity and Psychological Functioning in Children and Adolescents with Fontan Circulation

**DOI:** 10.1101/2024.04.16.24305934

**Authors:** Nicholas P. Seivert, Kathryn M. Dodds, Shannon O’Malley, David J. Goldberg, Stephen Paridon, Michael McBride, Jack Rychik

**Author notes:** Corresponding Author Contact Information: Nicholas Seivert, PhD, Assistant Professor of Clinical Psychiatry, Perelman School of Medicine at the University of Pennsylvania Pediatric Psychologist, Department of Child & Adolescent Psychiatry & Behavioral Sciences & The Cardiac Center, Children’s Hospital of Philadelphia, Mailing Address: CHOP Hub for Clinical Collaboration 3500 Civic Center Blvd, Room 12-550, Philadelphia, PA 19104.

## Abstract

**Background:** Pediatric patients with Fontan circulation (FC) often have diminished exercise capacity. They are also at risk for psychological problems, including anxiety, depression, and inattention. The current study examines associations between exercise capacity and psychological functioning in a clinical sample of pediatric patients with FC.

**Methods:** A multidisciplinary team evaluated participants in a dedicated clinic for pediatric patients with FC. Patients completed cardiopulmonary exercise testing (CPET); metrics selected for analyses included peak oxygen consumption (VO2), maximum achieved heart rate (HR), oxygen saturation at peak exercise, ventilatory anaerobic threshold, work rate, respiratory exchange ratio, and HR recovery at 1-, 3-, and 8-minutes after exercise. Parent and child were administered a standardized questionnaire to measure depression, anxiety, and inattention symptoms. Patients who completed CPET with adequate effort and a psychological measure were eligible for inclusion. Clinical data were extracted from medical records and analyzed using Pearson correlations.

**Results:** Clinical sample (n = 51) was 55% male with a mean age of 13.6 years (SD=2.5). A majority had hypoplastic left heart syndrome (51%). There was a significant negative correlation between parent-report of inattention and peak VO2 (*R*= -.307, 95% CI -.549/-.018, *P*= 0.038). Self-report of anxiety was positively correlated with HR recovery at 3 (*R*= .438, 95% CI .155/.655, *P*= 0.004) and 8 (*R*= .432, 95% CI .147/.651, *P*= 0.004) minutes post exercise. Depression was positively correlated with HR recovery at 3 minutes for parent report (*R*= .294, 95% CI .004/.538, *P*= 0.047) and 8 minutes for self-report (*R*= .410, 95% CI .122/.635, *P*= 0.007).

**Conclusions:** Inattention, depression, and anxiety symptoms were correlated with some CPET metrics in our clinical sample of pediatric patients with FC. Greater inattention symptoms were associated with lower peak VO2. Those with greater depression and anxiety symptoms showed a more rapid decline from max HR to recovery period.

**What is Known:**

- Many patients with Fontan circulation (FC) have some degree of diminished exercise capacity.
- The risk for mental health problems in pediatric patients with FC is very high, with an estimated lifetime prevalence of 65%.

**What the Study Adds:**

- There may be a link between mental health symptoms and exercise capacity metrics obtained during CPET for children and adolescents with FC.
- Our study found that greater inattention symptoms were linked to lower observed peak VO2 during CPET.
- Participants with higher levels of depression and anxiety symptoms exhibited a more dramatic decline in HR after exercise, contrary to findings in other cardiac patient samples that report an attenuated HR response for those with depression and anxiety.

## Introduction

Individuals with single ventricle congenital heart disease (CHD) have shown increased survival rates over the last few decades due to improvements in medical and surgical evaluation and management, including staged palliation culminating in the Fontan procedure.^1^ Despite these advances, individuals with Fontan circulation (FC) are at high risk for complications involving several organ systems.^2,3^ The FC impacts both physical and psychological functioning, and clinical surveillance recommendations have been developed for assessment and treatment of common concerns.^1^

Diminished exercise capacity is seen in individuals with FC and attributed to the cardiac physiology of chronically elevated central venous pressure and low cardiac output.^4^ Serial cardiopulmonary exercise testing (CPET) is an important method for assessing exercise capacity^1^ and also provides prognostic indicators of later complications in FC.^5^ As a group, individuals with FC tend to demonstrate poorer functioning compared to healthy subjects on several CPET metrics.^6^ Clinical characteristics of patients with FC associated with a key measure of physical fitness, higher peak oxygen consumption (peak VO2), include younger age, normal BMI, and healthier end organ functioning as reflected in laboratory values for musculoskeletal (vitamin D), hematologic (absolute lymphocyte count, hemoglobin), and hepatic (GGT, platelet count) systems.^7^

Autonomic dysfunction may be an important factor in understanding diminished exercise capacity in the FC. Mechanisms that may contribute to the diminished parasympathetic and sympathetic nervous system activation include surgery-related damage/denervation and specific hemodynamics characterized by low cardiac output.^8^ Heart rate (HR) recovery, the rate at which HR declines after exercise, is one metric of autonomic function and a prognostic indicator of morbidity and mortality in individuals with heart disease.^9^ Patients with FC typically demonstrate a blunted or delayed HR recovery after cessation of exercise during CPET.^10^

In addition to these physical challenges, individuals with FC are at increased risk for psychological and neurodevelopmental problems. Patients with FC have very high rates of mental health concerns, with estimates of up to 65% lifetime prevalence of psychiatric disorders, most commonly related to inattention and executive dysfunction, anxiety, and depression.^11,12^ Despite the increased risk for diminished exercise capacity, autonomic dysfunction, and psychological problems in youth with FC, there is little information on the potential relations among these variables in this unique population.

This study explores whether there are associations between physiologic measurements/parameters of exercise performance and psychological health in a sample of children and adolescents with FC. We hypothesized that inattention, anxiety, and depression symptoms would be associated with poorer exercise test performance. We also specifically examined whether greater anxiety and depression would be associated with attenuated HR recovery following exercise, reflecting autonomic dysfunction.

## Methods

Participants were a clinical sample of pediatric patients with single ventricle congenital heart disease and Fontan circulation. Patients were evaluated by the Children’s Hospital of Philadelphia FORWARD (Fontan Rehabilitation, Wellness, and Resilience Development) Clinic - a multidisciplinary team, including pediatric cardiology, immunology, endocrinology, gastroenterology, psychology, exercise physiology, nutrition, and social work. This dedicated clinic provides specialized surveillance for children and adolescents with Fontan circulation.^13^ Clinical and demographic data were extracted from patient medical records. This retrospective review was approved the Children’s Hospital of Philadelphia IRB.

As part of the evaluation, patients undergo cardiopulmonary exercise testing (CPET) primarily using a standard ramp cycle protocol.^7^ Exercise capacity metrics selected for analyses included peak VO2 (ml/kg/min), maximum HR achieved during exercise (max HR), oxygen saturation at maximum exercise (max O2 sat), oxygen consumption at ventilatory anaerobic threshold (VAT), maximum achieved work rate (WR), respiratory exchange ratio (RER), and HR recovery (max HR – post HR) at 1-, 3-, and 8-minutes following cessation of exercise.

The program psychologist completed an evaluation with all patients, including administration of standardized psychological screening measures completed by patients and their parents. The screening measure was the Behavioral Assessment System for Children, Third Edition (BASC-3), which provides a broad assessment of psychological functioning across several domains and includes both self-report and parent-report versions.^14^ Scales selected from the BASC-3 for analyses were based on common concerns seen in youth with FC as well as prior research showing associations between mental health symptoms and exercise capacity in other cardiac samples, including depression, anxiety, and attention problems. Scale scores are calculated using T-scores (mean=50, SD=10) comparing responses to a representative, norm-referenced sample based on child age.

Statistical analyses were conducted using IBM SPSS Statistics, Version 28. Analytic plan primarily involved calculation of Pearson correlations between mental health and CPET variables. Correlations selected for testing were based on hypotheses grounded in literature review. For mental health variables, separate analyses were conducted for child-report and parent-report of symptoms.

## Results

A total of 158 patients were evaluated in the FORWARD Program between January 2019 and June 2022. Data from a subsample (n=80) who completed the CPET ramp cycle protocol were eligible for inclusion in the study. The remaining patients either completed a different type of exercise protocol, including treadmill (n=31) and 6-minute walk (n=37), or had no data available (n=10). In the exercise lab, patients who complete protocols other than the ramp cycle are typically younger and unable to use the bike due to height. Additionally, a portion of the study period was at the height of the COVID-19 pandemic during which time safety precautions resulted in some inconsistency with patient completion of standard exercise testing.

Of the 80 patients who completed the CPET ramp cycle protocol, 9 displayed submaximal effort (RER < 1.10) and were excluded from analyses. Also excluded were patients on betablockers (n=1) and those with pacemakers (n=3) due to their influence on CPET variables of interest. This left a total of 67 patients from which psychological measures (BASC-3) were available for 51 patients, which were included in analyses for the current study. In terms of respondents for the psychological measure, 41 patients had both parent- and child-report, 7 had parent-only report, and 3 had child-only report. Statistical analyses comparing clinical and demographic characteristics of those with BASC-3 data and those without revealed no significant differences, with the exception of insurance status. Those participants who had commercial insurance were significantly more likely to have completed a BASC-3 measure compared to those with Medicaid or other/unknown insurance status (*χ*^2^= 11.96, P < 0.001).

Demographic and clinical characteristics are presented in Tables 1 and 2. A slight majority of patients were male, most identified their race/ethnicity as White/non-Hispanic/Latino, and patients primarily had private/commercial insurance. Just over half of the sample had a primary cardiac diagnosis of hypoplastic left heart syndrome. Mean age at the time of evaluation was 13.6 years (SD=2.5). Overall BMI of patients was within the normal range and mean height was slightly lower than expected for their age. Mean CPET metrics were similar to studies of individuals with FC in this age range, although oxygen saturation at maximal exercise was slightly lower in the current sample compared to others.^6,15^ Mean scores on the psychological measures (depression, anxiety, inattention) fell within the average range and were not indicative of difficulties in these domains for the sample overall.

**Table 1.** Clinical and Demographic Description of Sample.

| **Variable** |  | **N (% of Sample)** |
| --- | --- | --- |
| Sex | Male | 28 (55) |
|  | Female | 23 (45) |
| Race | White | 45 (88) |
|  | Black/African American | 1 (2) |
|  | Asian | 1 (2) |
|  | Other/unknown | 4 (8) |
| Ethnicity | Non-Hispanic/Latino | 47 (92) |
|  | Hispanic/Latino | 4 (8) |
| Insurance Status | Private/commercial | 42 (82) |
|  | Medicaid | 7 (14) |
|  | Other/unknown | 2 (4) |
| Primary Cardiac Diagnosis | Hypoplastic left heart syndrome | 26 (51) |
|  | Double inlet left ventricle | 5 (10) |
|  | Other single ventricle CHD | 20 (39) |
| **Total N** |  | **51 (100)** |

**Table 2.**
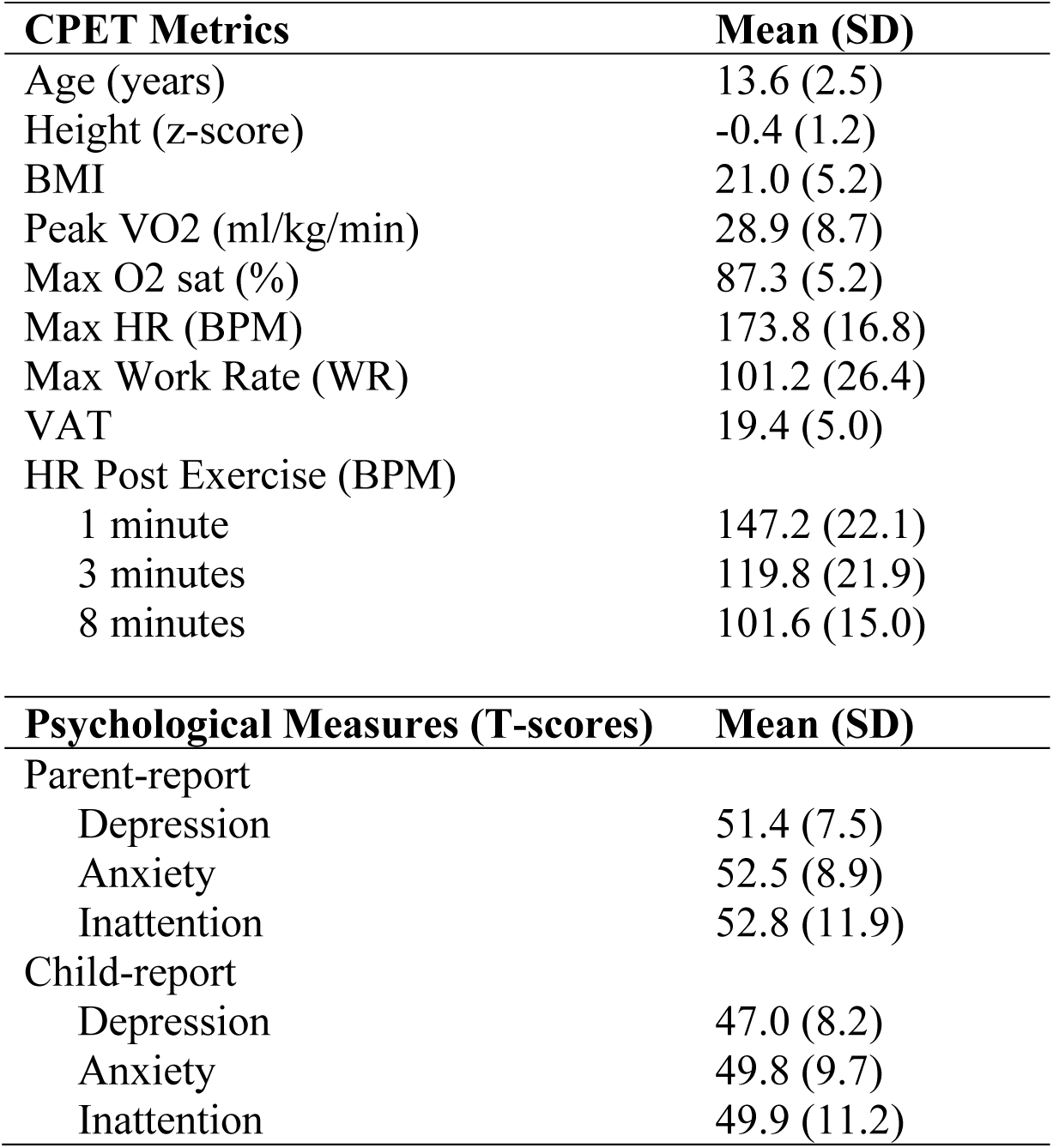
Sample Means and Standard Deviations of CPET and Psychological Measures.

| **CPET Metrics** | **Mean (SD)** |
| --- | --- |
| Age (years) | 13.6 (2.5) |
| Height (z-score) | -0.4 (1.2) |
| BMI | 21.0 (5.2) |
| Peak VO<sub>2</sub> (ml/kg/min) | 28.9 (8.7) |
| Max O<sub>2</sub> sat (%) | 87.3 (5.2) |
| Max HR (BPM) | 173.8 (16.8) |
| Max Work Rate (WR) | 101.2 (26.4) |
| VAT | 19.4 (5.0) |
| HR Post Exercise (BPM) |  |
| 1 minute | 147.2 (22.1) |
| 3 minutes | 119.8 (21.9) |
| 8 minutes | 101.6 (15.0) |
| **Psychological Measures (T-scores)** | **Mean (SD)** |
| Parent-report |  |
| Depression | 51.4 (7.5) |
| Anxiety | 52.5 (8.9) |
| Inattention | 52.8 (11.9) |
| Child-report |  |
| Depression | 47.0 (8.2) |
| Anxiety | 49.8 (9.7) |
| Inattention | 49.9 (11.2) |

Table 3 presents results of statistically significant analyses of Pearson correlations between psychological and CPET variables. There was a significant inverse relationship between parent-report of inattention and peak VO2, indicating those with greater attention problems demonstrated lower levels of oxygen consumption at peak exercise. There were no associations between depression and anxiety scores and metrics obtained during exercise.

**Table 3.** Statistically Significant Correlations Between CPET and Psychological Variables.

| **Psychological Symptoms** | **CPET Metrics** | **R** | **95% CI** | **P** |
| --- | --- | --- | --- | --- |
| Inattention (parent-report) | Peak VO2 | -.307 | -.549/-.018 | 0.038 |
| Anxiety (self-report) | HR Recovery 3 minutes | .438 | .155/.655 | 0.004 |
|  | HR Recovery 8 minutes | .432 | .147/.651 | 0.004 |
| Depression (parent-report) | HR Recovery 3 minutes | .294 | .004/.538 | 0.047 |
| Depression (self-report) | HR Recovery 8 minutes | .410 | .122/.635 | 0.007 |

As for the post-exercise HR variables, there was a moderate positive correlation between self-reported anxiety scores and HR recovery at 3- and 8-minutes, suggesting those with higher levels of anxiety showed a greater decrease in HR from peak exercise to the recovery period. Similarly, depression scores were positively associated with HR recovery, with parent-report of depression symptoms associated with HR recovery at 3-minutes and self-report with HR recovery at 8-minutes, indicating those with greater levels of depression showed a larger decline between peak and post-exercise HR.

## Discussion

The current study examined associations between psychological symptoms and exercise capacity metrics in a modest size sample of children and adolescents with FC. We found that attention problems, depression, and anxiety were correlated with some metrics obtained during CPET. Greater levels of parent-reported inattention symptoms were associated with lower peak VO2. We further found that those with greater levels of depression and anxiety symptoms showed a more rapid decline from max HR to recovery period HR at 3- and 8-minutes following cessation of exercise.

Executive dysfunction, including attention problems, can impact various domains of functioning. Relatedly, individuals with FC are at high risk for psychiatric disorders including attention deficit hyperactivity disorder (ADHD).^11^ Although the current sample was limited to those who displayed adequate effort, it is possible that those with greater levels of inattention had difficulty fully engaging with the exercise testing and managing the various demands placed on them during the task. Relatedly, there is some evidence from other research that greater levels of ADHD symptoms are associated with lower levels of cardiovascular fitness in the general pediatric population.^16^ Consistent with this, a small study of pediatric patients with FC examining neurodevelopmental correlates of CPET performance found parent ratings of adaptive functioning (e.g., activities of daily living, functional communication) were associated with higher peak VO2. In addition, these researchers found that those achieving higher HR during CPET demonstrated fewer attention problems on a performance measure of sustained visual attention.^17^ Inattention and executive dysfunction in those with FC clearly have the potential to impact exercise capacity metrics, however, specificity in terms what aspects of physical fitness these symptoms influence requires further study.

Our findings that greater depression and anxiety symptoms were associated with a steeper HR decline from exercise to recovery period were contrary to our expectations. As previously noted, HR recovery following cessation of exercise is an indicator of autonomic function. There is evidence in pediatric samples without cardiovascular disease that individuals with anxiety and depressive disorders demonstrate disrupted sympathetic and parasympathetic activation. A meta-analysis examining 20 studies of children and adolescents with clinical anxiety and depression found significantly lower heart rate variability, which suggests abnormal autonomic nervous system functioning.^18^ In addition, studies in adults with cardiac disease, including chronic heart failure^19^ and coronary heart disease^20^, found those who reported greater depression symptoms showed a blunted HR response at 1 minute after cessation of exercise. However, our current study subjects with FC exhibited the opposite effect with a more dramatic decline in HR, as opposed to an attenuation, being associated with higher scores on measures depression and anxiety. From the psychology perspective, our findings could be related to the distinction between state vs. trait anxiety. Trait anxiety is a personality characteristic associated with a genetic and biological predisposition towards perceiving threats in one’s environment, which is primarily what the current study’s anxiety measure assessed. State anxiety is related to the transitory response to specific environmental stressors^21^ and individuals with anxiety and depressive disorders are prone to high levels of both state and trait anxiety.^22^ In the current study, the relatively steeper decline in HR recovery for those with greater levels of anxiety and depression could be explained by a reduction of state anxiety in response to the acute removal of the environmental stressor of the exercise test. However, it is not entirely clear why this would be observed in our sample and not in other studies. Perhaps there is an age-related effect as the previously noted study finding a blunted HR response for those with anxiety/depression was with a sample of adults. In addition, there could be an interaction effect between anxiety/depression and the unique aspects of FC physiology in the single ventricle population resulting in the comparatively steeper decline in HR after cessation of exercise. Subjects in our study all underwent cardiac surgery, with the high likelihood for potential interruption of autonomic innervation of the heart. In particular, individuals with hypoplastic left heart syndrome, which made up a substantial portion of our cohort, all undergo extensive aortic surgical reconstruction, precisely in the region where autonomic cardiac innervation is located. Either inherent congenital differences in innervation from normal hearts in those with single ventricle, or mechanical disruption of nerve fibers at the time of surgery may lead to autonomic dysregulation and our findings.

Future research on links between anxiety/depression symptoms in FC and exercise capacity could attempt to differentiate between trait anxiety, which is more closely related to what the anxiety scale in our study measured, and state anxiety in the context of exercise testing. Additionally, alternative methods for measuring autonomic dysfunction, such as heart rate variability, and their associations with depression, anxiety, and exercise test performance could provide greater understanding of the relationship between psychological and physiological metrics in FC. Furthermore, assessing varying types of single ventricle congenital heart disease and factors involved in the perioperative course may identify distinguishing features that are either inherent in the congenital malformation or acquired as a consequence of the surgical interventions necessary. These more sophisticated analyses would require a larger sample than the current study. In terms of potential relations between inattention/executive dysfunction and exercise metrics, it may be helpful to have additional methods for assessing executive dysfunction, aside from just report of inattention symptoms, including neuropsychological measures. Furthermore, including a larger cohort may reveal a greater number of subjects manifesting abnormal and clinically significant levels of anxiety or depression which could allow for comparisons between this group and those without these concerns. Lastly, greater efforts are needed to include individuals with more diverse racial, ethnic, and socioeconomic backgrounds to increase generalizability of findings and determine any potential impact of these factors on outcomes.

Despite historical activity restrictions on physical activity for these patients, evidence increasingly supports that exercise is beneficial to individuals with Fontan circulation.^23^ Although there is increased risk of both diminished exercise capacity and mental health issues, limited prior research has examined whether there are associations between aspects of psychosocial functioning and physical fitness in FC. The current study contributes to a growing literature on interrelation of medical and psychological concerns often observed in this populations. It also highlights the importance of including psychologists and other mental health professionals into the standard of care for all patients with FC in order to provide truly comprehensive healthcare services tailored to the unique needs of these individuals.

## Data Availability

Clinical data was extracted from patient medical records and is stored securely by the first author.

## Acknowledgements

None

## Sources of Funding

None

## Disclosures

None

## Notes

### Competing Interest Statement

The authors have declared no competing interest.

### Clinical Trial

N/a

### Author Declarations

Research was approved by the Children's Hospital of Philadelphia IRB.

